# Comparison of cardiovascular and renal outcomes between dapagliflozin and empagliflozin in patients with type 2 diabetes

**DOI:** 10.1101/2022.05.23.22275457

**Authors:** Jayoung Lim, In-Chang Hwang, Hong-Mi Choi, Yeonyee E. Yoon, Goo-Yeong Cho

**Affiliations:** Department of Cardiology, Cardiovascular Center, Seoul National University Bundang Hospital, Seongnam, Gyeonggi, South Korea; Department of Internal Medicine, Seoul National University College of Medicine, Seoul, South Korea

## Abstract

**Background:** The cardiovascular and renal benefits of sodium glucose co-transporter 2 inhibitors (SGLT2i) have been clearly demonstrated. However, studies comparing the effects of dapagliflozin and empagliflozin are scarce. Therefore, we aimed to compare the clinical outcomes between dapagliflozin and empagliflozin in patients with type 2 diabetes without prior atherosclerotic cardiovascular disease, chronic kidney disease, or heart failure.

**Methods:** Using a propensity-score matching method, we retrospectively analyzed 921 patients treated with dapagliflozin, 921 patients treated with empagliflozin, and 1842 patients treated with dipeptidyl peptidase-4 inhibitors (DPP4i; control group). Study outcomes comprised composite coronary events (acute coronary syndrome and coronary revascularization), composite ischemic events (coronary events and stroke), and composite heart failure and renal events.

**Results:** During follow up (median, 43.4 months), the incidence of composite coronary events was significantly lower in the SGLT2i groups than in the control group, and the incidence of composite ischemic events was lower in the dapagliflozin group than in the control group. Dapagliflozin and empagliflozin both demonstrated significant benefits in terms of heart failure and renal outcomes, supported by renoprotective effects, as assessed by the change in glomerular filtration rate. At 24–36 months of treatment, the empagliflozin group had higher low-density lipoprotein cholesterol levels, and lower glycated hemoglobin levels, compared to those in the dapagliflozin and control groups.

**Conclusion:** SGLT2i use was associated with a significantly reduced risk of atherosclerotic cardiovascular events, heart failure hospitalization, and renal events, compared to that with DPP4i use. There were no significant differences in clinical outcomes between dapagliflozin and empagliflozin, supporting a SGLT2i class effect.

## Introduction

Type 2 diabetes is a major risk factor for macrovascular and microvascular diseases (1). The treatment strategy for type 2 diabetes has evolved from glycemic control to patient-centered approaches, with consideration of the risk of atherosclerotic cardiovascular disease (ASCVD), chronic kidney disease (CKD), and heart failure (HF) (2). This expansion in therapeutic strategy is mainly based on the introduction of new anti-hyperglycemic agents, such as sodium glucose co-transporter 2 inhibitors (SGLT2i), which were the first to demonstrate a prognostic benefit. Large-scale clinical trials of two SGLT2i, empagliflozin and dapagliflozin, have shown direct and indirect evidence for their significant cardiovascular and renal protective effects in patients with type 2 diabetes (3-8). As microvascular dysfunction, inflammation, oxidative stress, and fibrosis contribute to the progression of diabetic kidney disease and HF, SGLT2i-mediated attenuations in these pathways may contribute to cardiovascular and renal protection (9, 10). Several meta-analyses have suggested that there is a class effect for SGLT2i (11-14). However, a few retrospective studies have suggested differences in effects between SGLT2i, favoring the use of dapagliflozin over empagliflozin in terms of HF prevention (15, 16). On the other hand, a few studies have indicated more favorable glycemic control and management of cardiometabolic parameters with empagliflozin than with dapagliflozin (17, 18). Despite potential differences in benefit profiles, no trial has directly compared these SGLT2i in terms of clinical outcomes.

Therefore, in the present retrospective study, we aimed to compare the cardiovascular and renal outcomes between dapagliflozin and empagliflozin in patients with type 2 diabetes without prior ASCVD, CKD, or HF, and assess their impact on lipid profiles, glycemic control, and renal function.

## Materials and methods

### Study population

Patients with type 2 diabetes prescribed empagliflozin, dapagliflozin, or a dipeptidyl peptidase-4 inhibitor (DPP4i) at Seoul National University Bundang Hospital from April 2009 to December 2020 were retrospectively identified (19). The date of medication (SGLT2i or DPP4i) initiation was defined as the index date. Exclusion criteria were type 1 diabetes mellitus, prior HF, prior ASCVD (angina, myocardial infarction, coronary revascularization, peripheral artery disease, and stroke), prior CKD (glomerular filtration rate [GFR] <45 mL/min/1.73m^2^), short duration of medication use (<3 months), low medication possession ratio (<75%), and simultaneous or sequential use of empagliflozin and dapagliflozin.

In total, 921 patients treated with empagliflozin, 1,424 patients treated with dapagliflozin, and 10,981 patients treated with DPP4i (control group) were identified (**Fig 1**). After propensity-score matching (1:1:2 ratio) on clinical factors, laboratory findings, and medication use, the empagliflozin and dapagliflozin groups comprised 921 patients each, and the DPP4i (control) group comprised 1,842 patients (**Fig 1**). Seoul National Bundang Hospital’s Institutional Review Board approved the study protocol and waived the requirement for informed consent.

**Fig 1.**
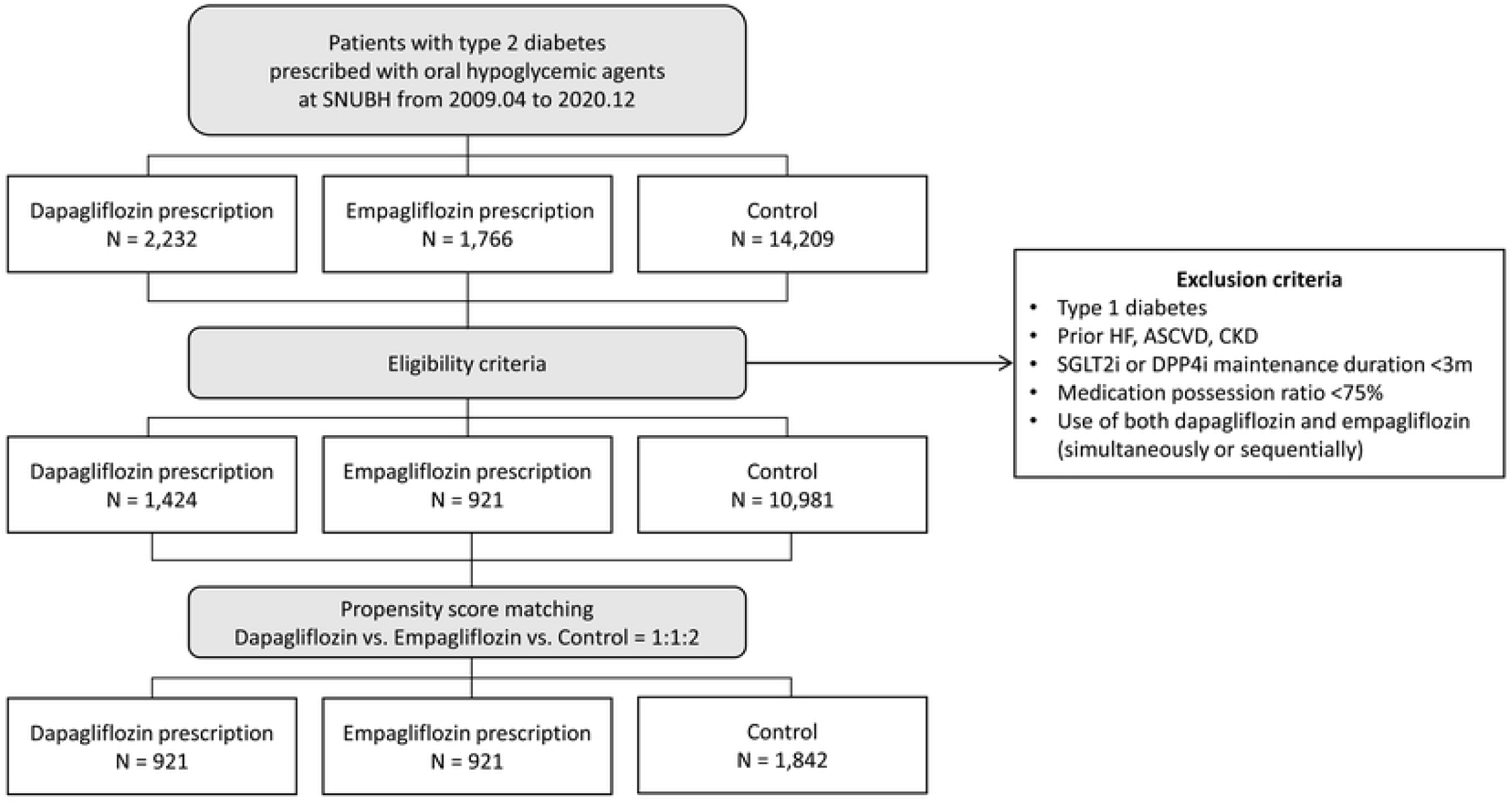
Study flowchart. After 1:1:2 propensity score matching, there were 921 patients treated with empagliflozin, 921 patients treated with dapagliflozin, and 1,842 patients treated with DPP4i (control group). ASCVD, atherosclerotic cardiovascular disease; CKD, chronic kidney disease; DPP4i, dipeptidyl peptidase-4 inhibitor; GLP1RA, glucagon-like peptide 1 receptor agonists; HF, heart failure; SGLT2i, sodium-glucose co-transporter 2 inhibitor

### Measurements

We collected data on age, sex, systolic blood pressure, diastolic blood pressure, history of smoking, preexisting comorbidities, medications, and laboratory findings, including the lipid profile, fasting glucose level, glycated hemoglobin (HbA1c) level, and GFR.

### Outcomes

Study outcomes comprised composite coronary events (acute coronary syndrome and coronary revascularization), composite ischemic events (acute coronary syndrome, coronary revascularization, and stroke), hospitalization for HF (HHF), renal events (renal death, initiation of renal replacement therapy, and admission due to acute kidney injury or CKD progression), and composite HHF and renal events. Changes in the levels of total cholesterol, low-density lipoprotein cholesterol (LDLc), and HbA1c, and GFR during follow up were also assessed.

### Statistical analysis

To adjust for imbalances in the baseline characteristics of patients among empagliflozin, dapagliflozin, and control groups, propensity-score matching, with a 1:1:2 ratio, was performed using the nearest neighbor method, with the following covariates: age, sex, 10-year ASCVD risk, systolic blood pressure, presence of microalbuminuria/proteinuria, HbA1c level, GFR, and the use of antiplatelet agents, renin-angiotensin system blockers, and statins. The distributions of propensity scores and standardized mean differences were calculated to assess the strength of matching.

Categorical variables are presented as numbers with percentages, and continuous variables as medians with interquartile ranges (IQRs). Group comparisons were performed using the χ2 test for categorical variables and analysis of variance for continuous variables. Hazard ratios (HRs) with 95% confidence intervals (CIs) were estimated using the Cox proportional-hazards method.

Statistical analyses were performed using SPSS version 22.0 (IBM Co., Armonk, NY, USA) and R version 4.0.2 (The R Foundation for Statistical Computing, Vienna, Austria). Two-sided p-values <0.05 were considered statistically significant.

## Results

### Baseline characteristics

The baseline characteristics, laboratory findings, and use of medications were well balanced among the groups (**Table 1**). The median age was 56, 56, and 57 years in the dapagliflozin, empagliflozin, and control groups, respectively, and two-thirds of the patients were male. The prevalence of current smoking, hypertension, and dyslipidemia was 20%, 60%, and 80%, respectively. The mean 10-year ASCVD risk ranged 10%–11%, without significant inter-group differences. In all three groups, the median HbA1c level was approximately 8.0% and median GFR was approximately 97 mL/min/1.73m^2^, and the prevalence of microalbuminuria or proteinuria was more than 30%. Two-thirds of the patients were on statins, and less than 50% were on RAS blockers. Regarding anti-diabetic medication, more than 97% of the patients were on metformin, more than 40% were on sulfonylurea, and 18% were on insulin therapy.

**Table 1.**
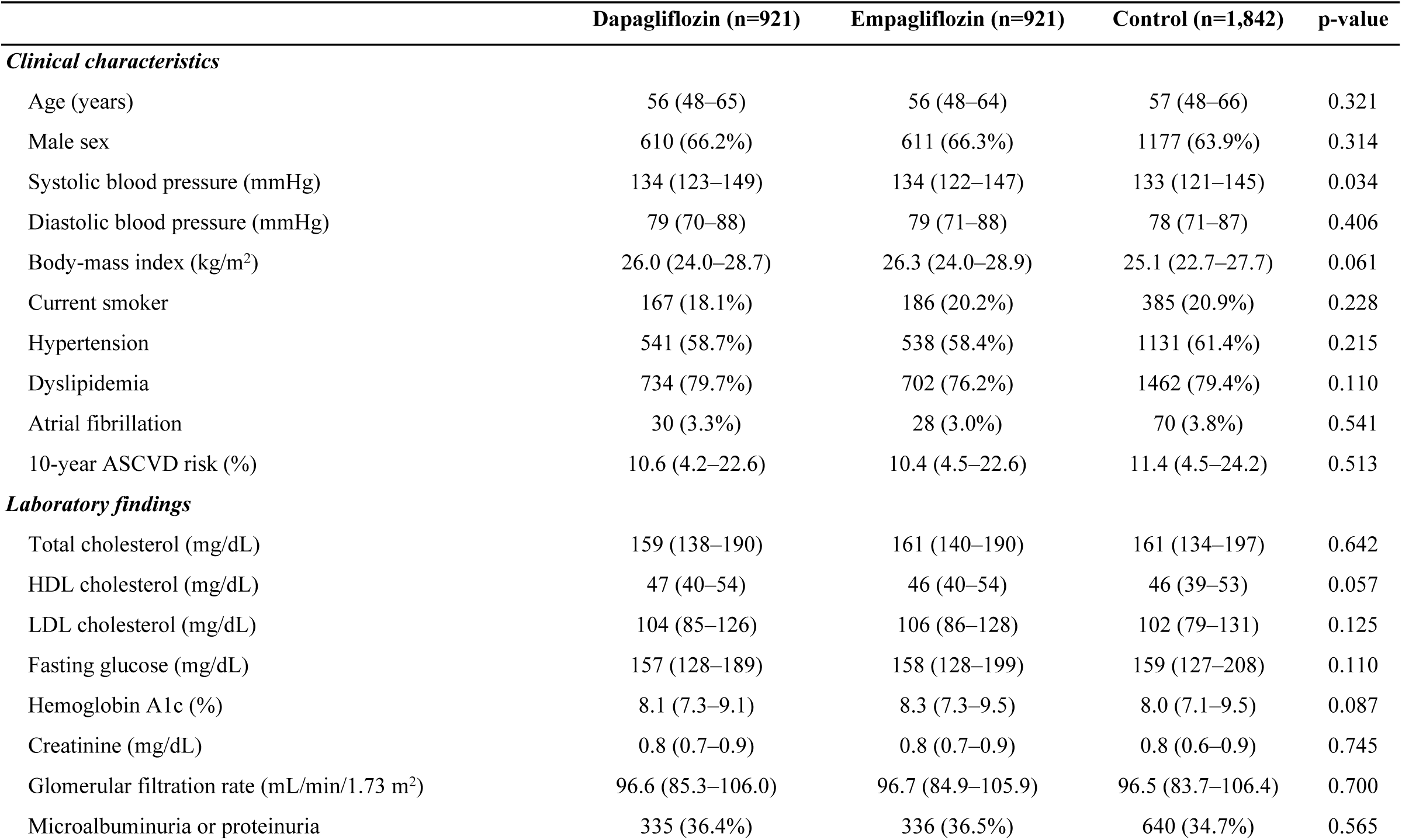
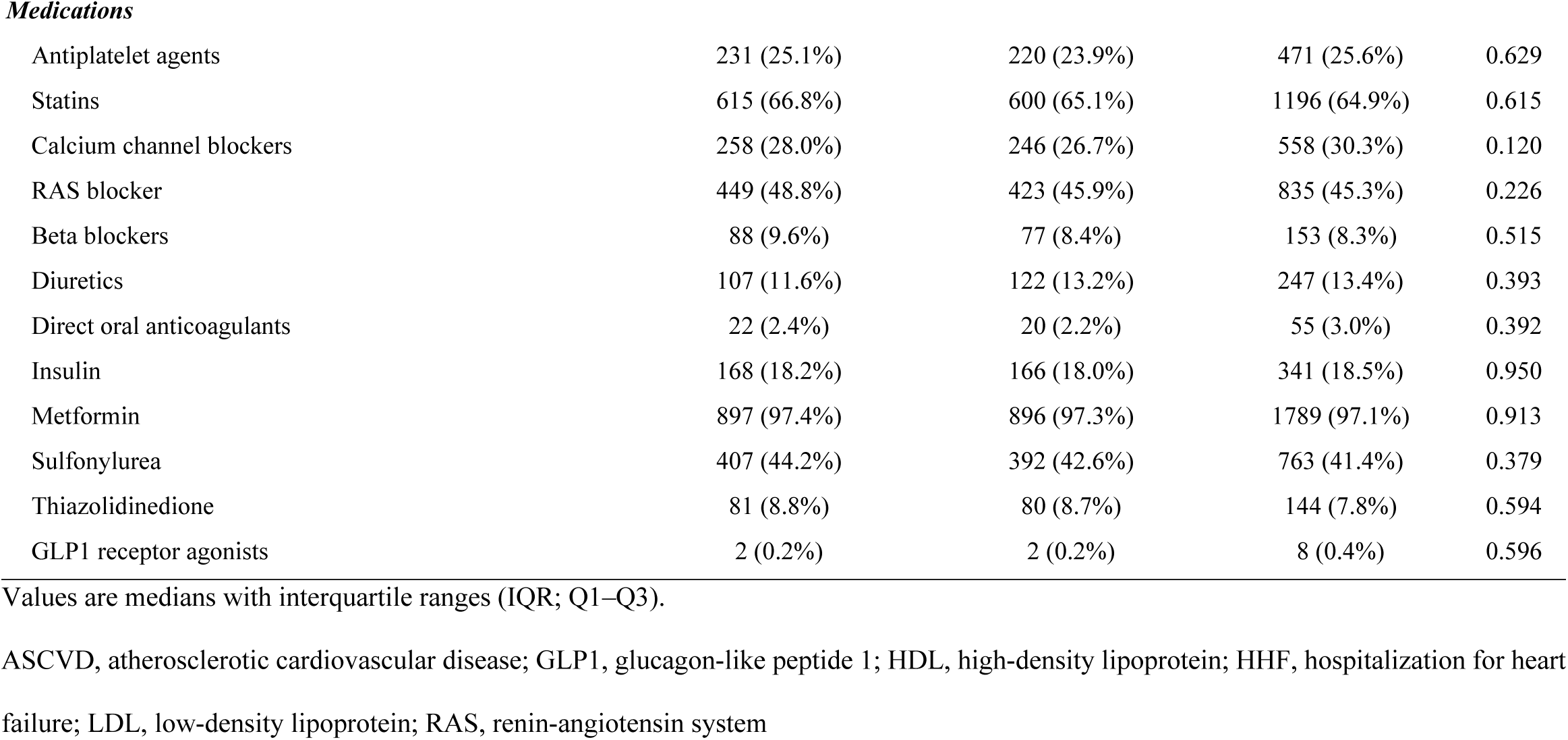
Baseline characteristics.

|  | Dapagliflozin (n=921) | Empagliflozin (n=921) | Control (n=1,842) | p-value |
| --- | --- | --- | --- | --- |
| ***Clinical characteristics*** |  |  |  |  |
| Age (years) | 56 (48–65) | 56 (48–64) | 57 (48–66) | 0.321 |
| Male sex | 610 (66.2%) | 611 (66.3%) | 1177 (63.9%) | 0.314 |
| Systolic blood pressure (mmHg) | 134 (123–149) | 134 (122–147) | 133 (121–145) | 0.034 |
| Diastolic blood pressure (mmHg) | 79 (70–88) | 79 (71–88) | 78 (71–87) | 0.406 |
| Body-mass index (kg/m<sup>2</sup>) | 26.0 (24.0–28.7) | 26.3 (24.0–28.9) | 25.1 (22.7–27.7) | 0.061 |
| Current smoker | 167 (18.1%) | 186 (20.2%) | 385 (20.9%) | 0.228 |
| Hypertension | 541 (58.7%) | 538 (58.4%) | 1131 (61.4%) | 0.215 |
| Dyslipidemia | 734 (79.7%) | 702 (76.2%) | 1462 (79.4%) | 0.110 |
| Atrial fibrillation | 30 (3.3%) | 28 (3.0%) | 70 (3.8%) | 0.541 |
| 10-year ASCVD risk (%) | 10.6 (4.2–22.6) | 10.4 (4.5–22.6) | 11.4 (4.5–24.2) | 0.513 |
| ***Laboratory findings*** |  |  |  |  |
| Total cholesterol (mg/dL) | 159 (138–190) | 161 (140–190) | 161 (134–197) | 0.642 |
| HDL cholesterol (mg/dL) | 47 (40–54) | 46 (40–54) | 46 (39–53) | 0.057 |
| LDL cholesterol (mg/dL) | 104 (85–126) | 106 (86–128) | 102 (79–131) | 0.125 |
| Fasting glucose (mg/dL) | 157 (128–189) | 158 (128–199) | 159 (127–208) | 0.110 |
| Hemoglobin A1c (%) | 8.1 (7.3–9.1) | 8.3 (7.3–9.5) | 8.0 (7.1–9.5) | 0.087 |
| Creatinine (mg/dL) | 0.8 (0.7–0.9) | 0.8 (0.7–0.9) | 0.8 (0.6–0.9) | 0.745 |
| Glomerular filtration rate (mL/min/1.73 m<sup>2</sup>) | 96.6 (85.3–106.0) | 96.7 (84.9–105.9) | 96.5 (83.7–106.4) | 0.700 |
| Microalbuminuria or proteinuria | 335 (36.4%) | 336 (36.5%) | 640 (34.7%) | 0.565 |
| **Medications** |  |  |  |  |
| Antiplatelet agents | 231 (25.1%) | 220 (23.9%) | 471 (25.6%) | 0.629 |
| Statins | 615 (66.8%) | 600 (65.1%) | 1196 (64.9%) | 0.615 |
| Calcium channel blockers | 258 (28.0%) | 246 (26.7%) | 558 (30.3%) | 0.120 |
| RAS blocker | 449 (48.8%) | 423 (45.9%) | 835 (45.3%) | 0.226 |
| Beta blockers | 88 (9.6%) | 77 (8.4%) | 153 (8.3%) | 0.515 |
| Diuretics | 107 (11.6%) | 122 (13.2%) | 247 (13.4%) | 0.393 |
| Direct oral anticoagulants | 22 (2.4%) | 20 (2.2%) | 55 (3.0%) | 0.392 |
| Insulin | 168 (18.2%) | 166 (18.0%) | 341 (18.5%) | 0.950 |
| Metformin | 897 (97.4%) | 896 (97.3%) | 1789 (97.1%) | 0.913 |
| Sulfonylurea | 407 (44.2%) | 392 (42.6%) | 763 (41.4%) | 0.379 |
| Thiazolidinedione | 81 (8.8%) | 80 (8.7%) | 144 (7.8%) | 0.594 |
| GLP1 receptor agonists | 2 (0.2%) | 2 (0.2%) | 8 (0.4%) | 0.596 |

Values are medians with interquartile ranges (IQR; Q1–Q3).
ASCVD, atherosclerotic cardiovascular disease; GLP1, glucagon-like peptide 1; HDL, high-density lipoprotein; HHF, hospitalization for heart
failure; LDL, low-density lipoprotein; RAS, renin-angiotensin system

### Clinical outcomes

The median follow-up duration was 37.3 months (IQR, 24.5–55.7 months), 37.1 months (IQR 27.1–49.2 months), and 55.1 months (27.5–84.9 months) in the dapagliflozin, empagliflozin, and control groups, respectively (**Table 2**). Composite coronary events occurred in 6 patients (0.7%) in the dapagliflozin group, 10 patients (1.1%) in the empagliflozin group, and 55 patients (3.0%) in the control group. On multivariable analysis, advanced age, current smoking, presence of hypertension, and higher 10-year ASCVD risk were associated with a higher risk of composite coronary and ischemic events (**Table 3**). SGLT2i use was significantly associated with a lower incidence of composite coronary events (dapagliflozin vs. control: HR, 0.267; 95% CI, 0.114–0.627; p=0.002) (empagliflozin vs. control: HR, 0.467; 95% CI, 0.235– 0.929; p=0.030), without a significant difference between dapagliflozin and empagliflozin groups (HR, 2.196; 95% CI, 0.742–6.502; p=0.156) (**Fig 2A**). Likewise, compared to that in the control group, the occurrence of composite ischemic events was significantly less frequent in the dapagliflozin group, and tended to be lower in the empagliflozin group, without a significant difference between dapagliflozin and empagliflozin groups (**Fig 2B**).

**Table 2.** Clinical outcomes.

|  | **Dapagliflozin (n=921)** | **Empagliflozin (n=921)** | **Control (n=1,842)** | **p-value** |
| --- | --- | --- | --- | --- |
| Follow-up duration (months) | 37.3 (24.5–55.7) | 37.1 (27.1–49.2) | 55.1 (27.5–84.9) | <0.001 |
| All-cause death | 2 (0.2%) | 2 (0.2%) | 60 (3.3%) | <0.001 |
| Cardiovascular death | 1 (0.1%) | 2 (0.2%) | 5 (0.3%) | 0.897 |
| Acute coronary syndrome | 2 (0.2%) | 2 (0.2%) | 31 (1.7%) | <0.001 |
| Coronary revascularization | 5 (0.5%) | 7 (0.8%) | 39 (2.1%) | 0.001 |
| Stroke | 3 (0.3%) | 2 (0.2%) | 4 (0.2%) | 0.908 |
| HHF | 3 (0.3%) | 6 (0.7%) | 40 (2.2%) | <0.001 |
| Renal events (renal admission, progression to ESRD) | 2 (0.2%) | 2 (0.2%) | 30 (1.6%) | <0.001 |
| Composite coronary events | 6 (0.7%) | 10 (1.1%) | 55 (3.0%) | <0.001 |
| Composite ischemic events | 11 (1.2%) | 14 (1.5%) | 59 (3.2%) | 0.001 |
| Composite of HHF and renal events | 4 (0.4%) | 8 (0.9%) | 64 (3.5%) | <0.001 |

ESRD, end-stage renal disease; HHF, hospitalization for heart failure

**Table 3.**
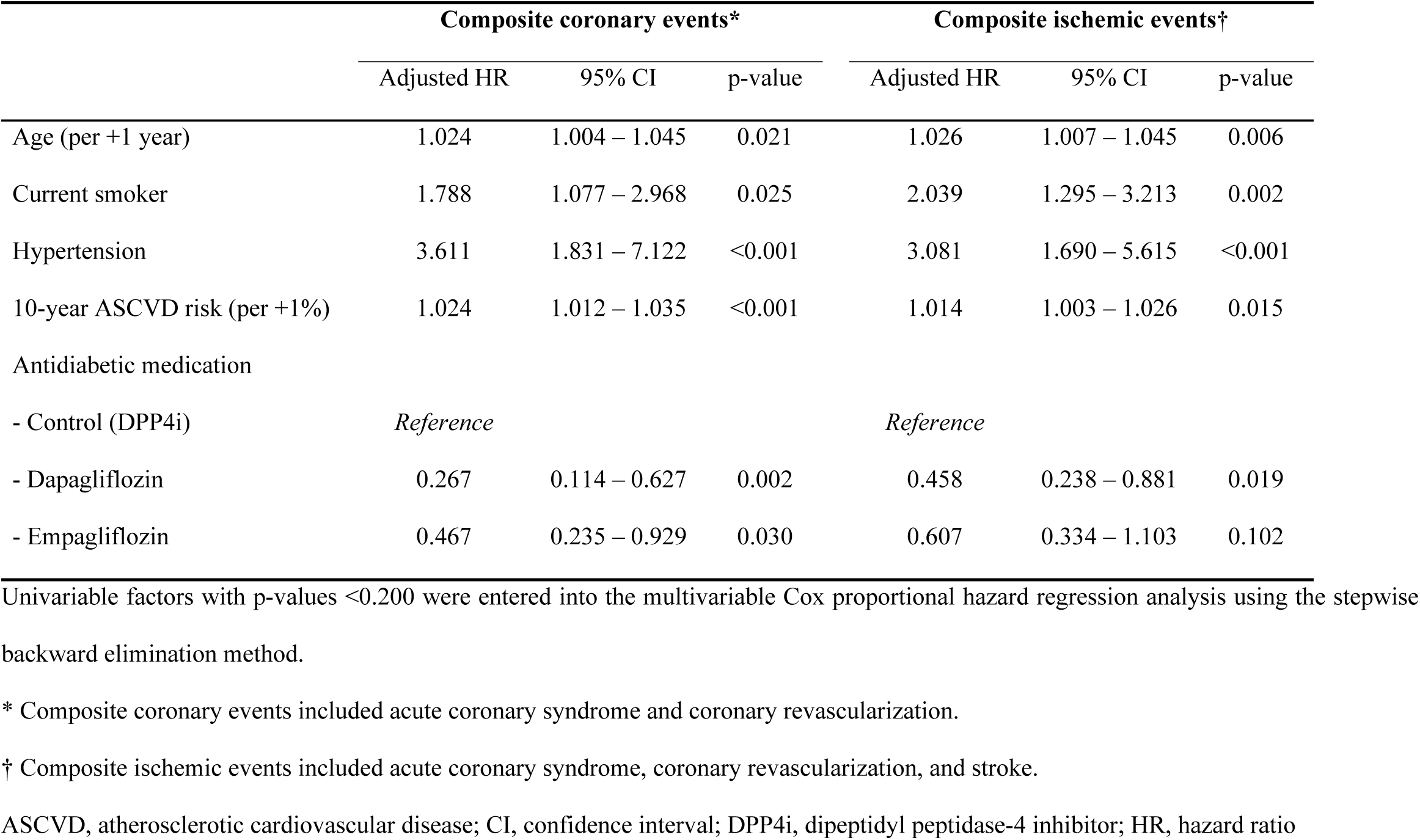
Multivariable predictors of ischemic events.

|  | Composite coronary events* |  |  | Composite ischemic events† |  |  |
| --- | --- | --- | --- | --- | --- | --- |
|  | Adjusted HR | 95% CI | p-value | Adjusted HR | 95% CI | p-value |
| Age (per +1 year) | 1.024 | 1.004 – 1.045 | 0.021 | 1.026 | 1.007 – 1.045 | 0.006 |
| Current smoker | 1.788 | 1.077 – 2.968 | 0.025 | 2.039 | 1.295 – 3.213 | 0.002 |
| Hypertension | 3.611 | 1.831 – 7.122 | <0.001 | 3.081 | 1.690 – 5.615 | <0.001 |
| 10-year ASCVD risk (per +1%) | 1.024 | 1.012 – 1.035 | <0.001 | 1.014 | 1.003 – 1.026 | 0.015 |
| Antidiabetic medication |  |  |  |  |  |  |
| - Control (DPP4i) | *Reference* |  |  | *Reference* |  |  |
| - Dapagliflozin | 0.267 | 0.114 – 0.627 | 0.002 | 0.458 | 0.238 – 0.881 | 0.019 |
| - Empagliflozin | 0.467 | 0.235 – 0.929 | 0.030 | 0.607 | 0.334 – 1.103 | 0.102 |

Univariable factors with p-values <0.200 were entered into the multivariable Cox proportional hazard regression analysis using the stepwise backward elimination method.
\* Composite coronary events included acute coronary syndrome and coronary revascularization.
† Composite ischemic events included acute coronary syndrome, coronary revascularization, and stroke.
ASCVD, atherosclerotic cardiovascular disease; CI, confidence interval; DPP4i, dipeptidyl peptidase-4 inhibitor; HR, hazard ratio

**Fig 2.**
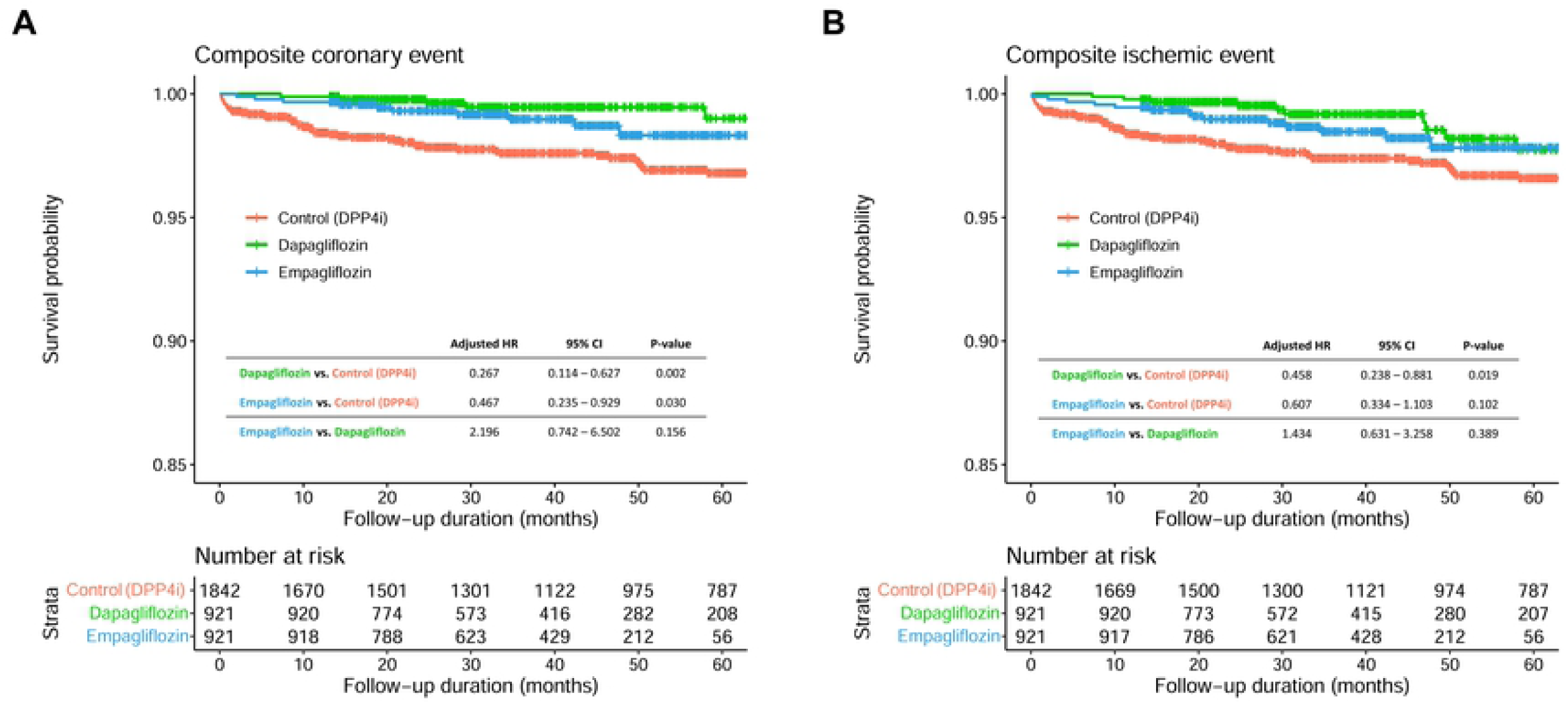
Event-free survival curves for atherosclerotic events. Kaplan-Meier curves comparing the risk of **(A)** composite coronary events and **(B)** composite ischemic events between the 3 groups. CI, confidence interval; HR, hazard ratio; SGLT2i, sodium-glucose co-transporter 2 inhibitor

Composite HHF and renal events occurred in 4 patients (0.4%) in the dapagliflozin group and 8 patients (0.9%) in the empagliflozin group (**Table 2**). Significant risk factors for HHF and renal events comprised atrial fibrillation, impaired renal function, and the presence of microalbuminuria or proteinuria (**Table 4**). Both dapagliflozin and empagliflozin demonstrated significant beneficial effects against HHF and renal events compared to that with DPP4i (dapagliflozin vs. control: HR, 0.186; 95% CI, 0.067–0.516; p=0.001) (empagliflozin vs. control: HR, 0.358; 95% CI, 0.169–0.756; p=0.007) (**Fig 3**).

**Table 4.** Multivariable predictors of HF and renal events.

|  | HHF |  |  | Renal events* |  |  | Composite of HHF and renal events |  |  |
| --- | --- | --- | --- | --- | --- | --- | --- | --- | --- |
|  | Adjusted HR | 95% CI | p-value | Adjusted HR | 95% CI | p-value | Adjusted HR | 95% CI | p-value |
| Age (per +1 year) | 1.022 | 0.998 – 1.048 | 0.076 | - | - | - | - | - | - |
| Hypertension | 1.961 | 0.960 – 4.003 | 0.064 | - | - | - | - | - | - |
| Atrial fibrillation | 6.888 | 3.398 – 13.961 | <0.001 | - | - | - | 4.431 | 2.355 – 8.337 | <0.001 |
| HbA1c (per +1%) | 1.145 | 1.002 – 1.309 | 0.047 | 0.786 | 0.622 – 0.994 | 0.044 | - | - | - |
| GFR (per +1 mL/min/1.73m<sup>2</sup>) | - | - | - | 0.971 | 0.954 – 0.988 | 0.001 | 0.983 | 0.972 – 0.995 | 0.005 |
| Microalbuminuria or proteinuria | 1.668 | 0.928 – 2.996 | 0.087 | 2.993 | 1.444 – 6.205 | 0.003 | 2.225 | 1.402 – 3.531 | 0.001 |
| Antidiabetic medication |  |  |  |  |  |  |  |  |  |
| - Control (DPP4i) | *Reference* |  |  | *Reference* |  |  | *Reference* |  |  |
| - Dapagliflozin | 0.182 | 0.056 – 0.593 | 0.005 | 0.235 | 0.055 – 1.002 | 0.050 | 0.186 | 0.067 – 0.516 | 0.001 |
| - Empagliflozin | 0.349 | 0.147 – 0.832 | 0.018 | 0.219 | 0.051 – 0.932 | 0.040 | 0.358 | 0.169 – 0.756 | 0.007 |

Univariable factors with p-values <0.200 were entered into the multivariable Cox proportional hazard regression analysis using the stepwise backward elimination method.
\* Renal events included renal death, hospitalization for acute kidney injury, and progression to end-stage renal disease.
CI, confidence interval; DPP4i, dipeptidyl peptidase-4 inhibitor; GFR, glomerular filtration rate; HbA1c, glycated hemoglobin A1c; HHF,

**Fig 3.**
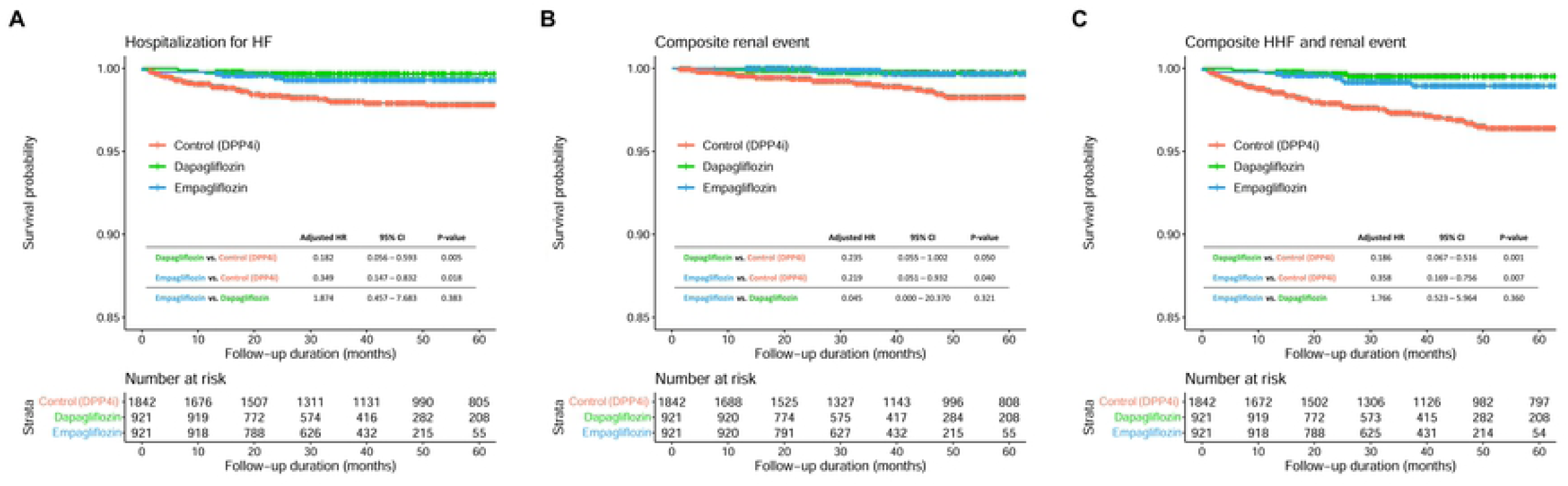
Event-free survival curves for HHF and renal events. Kaplan-Meier curves comparing the risk of **(A)** HHF, **(B)** composite renal events, and **(C)** composite HHF and renal events between the 3 groups. CI, confidence interval; HHF, hospitalization for heart failure; HR, hazard ratio; SGLT2i, sodium-glucose co-transporter 2 inhibitor

### Laboratory findings

Changes in the levels of total cholesterol, LDLc, and HbA1c, and GFR during follow up are shown according to group in **Fig 4**. At baseline, there were no significant inter-group differences in the levels of total cholesterol, LDLc, and HbA1c, and GFR. Overall, the total cholesterol level did not show inter-group differences during follow up (**Fig 4A**). However, at 24 and 36 months of treatment, the LDLc level was significantly higher in the empagliflozin group than in the control and dapagliflozin groups (**Fig 4B**), and the HbA1c level was significantly lower in the empagliflozin group than in the other groups (**Fig 4C**). Additionally, the GFR level tended to be better preserved at 24 months of treatment, and was significantly higher at 36 months and 48 months of treatment, in the dapagliflozin and empagliflozin groups than in the control group (**Fig 4D**).

**Fig 4.**
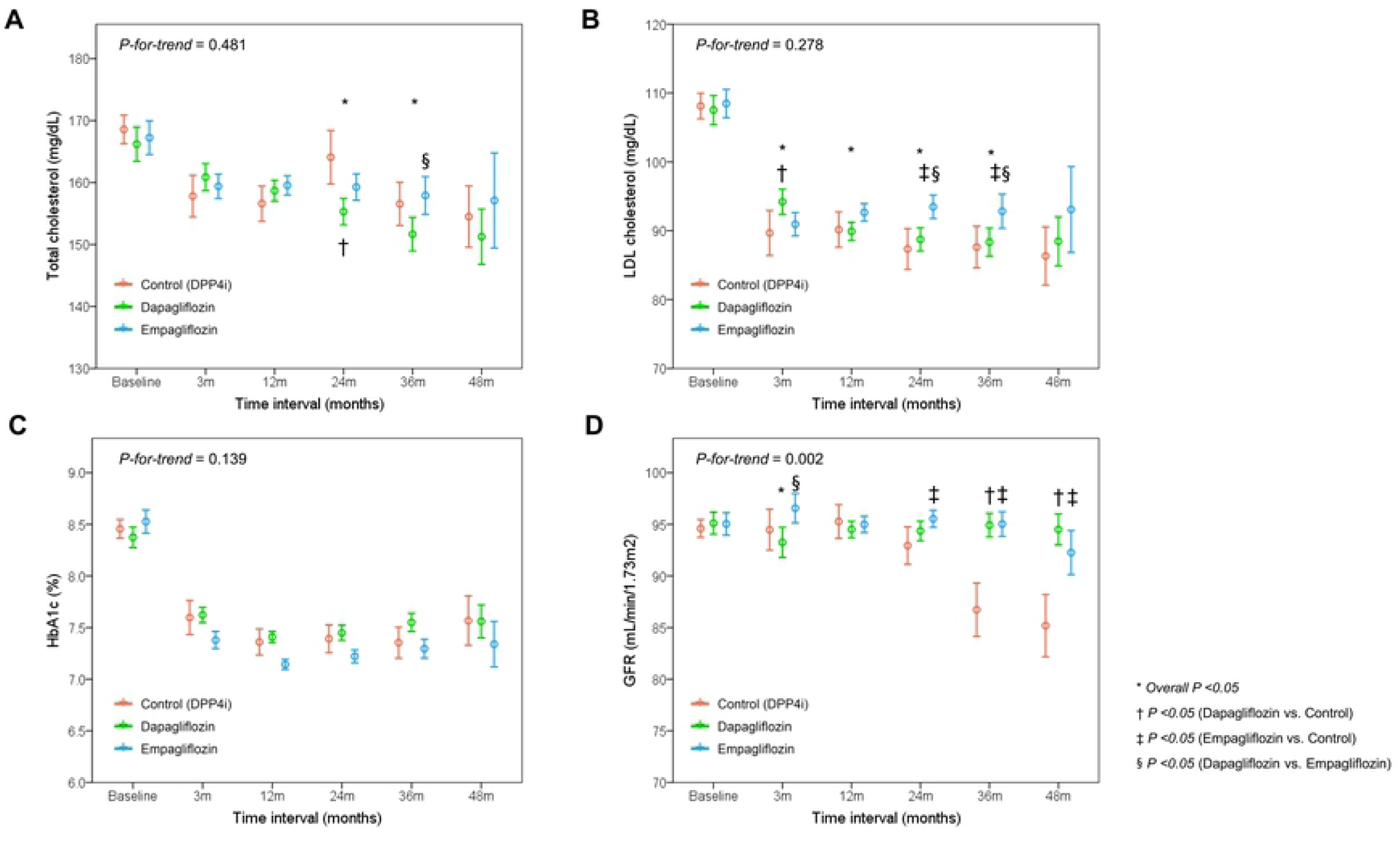
Changes in laboratory findings. Serial changes in **(A)** total cholesterol, **(B)** LDL cholesterol, **(C)** HbA1c, and **(D)** GFR are plotted according to group. GFR, glomerular filtration rate; HbA1c, glycosylated hemoglobin; LDL, low-density lipoprotein; SGLT2i, sodium-glucose co-transporter 2 inhibitor

## Discussion

In this retrospective study, we compared cardiovascular and renal outcomes among dapagliflozin, empagliflozin, and DPP4i (control) groups in 3,684 patients with type 2 diabetes and without prior ASCVD, HF, and CKD. Patients treated with SGLT2i demonstrated a significantly lower risk of atherosclerotic events, HHF, and renal events than that in patients treated with DPP4i (control group), with significant benefits in renal protection as assessed by the GFR level. There were no significant differences in clinical outcomes between dapagliflozin and empagliflozin groups. Of note, at 24–36 months of treatment, we observed higher LDLc levels and lower HbA1c levels in the empagliflozin group than in the other groups.

### Clinical benefits of SGLT2i

A series of landmark trials provided robust evidence on the cardiovascular and renal benefits of SGLT2i, regardless of baseline ASCVD status. In the EMPA-REG outcome trial, empagliflozin was associated with a reduced risk of cardiovascular death (HR, 0.86; 95% CI, 0.74 to 0.99; p=0.04 for superiority) in patients with type 2 diabetes and established ASCVD (6). Canagliflozin showed a reduced risk for HHF and cardiovascular death (HR, 0.78; 95% CI, 0.52-0.87; p=0.02) and composite renal outcomes (HR, 0.6; 95% CI, 0.47 to 0.77; p<0.001) in patients with type 2 diabetes, two-thirds of whom had established ASCVD (3, 4). In the DECLARE–TIMI 58 trial, dapagliflozin showed a lower rate of cardiovascular death and HHF compared to that with placebo in patients with type 2 diabetes with established ASCVD (40%) or at high risk for ASCVD (60%) (HR, 0.83; 95% CI, 0.73–0.95; p=0.005) (5). In a meta-analysis, SGLT2i use was associated with a reduced risk of major adverse cardiovascular events, HHF, and adverse kidney outcomes, independent of baseline ASCVD status (20).

Furthermore, the benefits of SGLT2i use have been observed in patients without prior cardiovascular disease, HF, or CKD. In the EMPRISE East Asia study, empagliflozin treatment was associated with a 28% reduced risk for HHF in patients with diabetes without CVD (21); SGLT2i use showed a 36% risk reduction for HHF among patients with diabetes without prior HF (22); and SGLT2i use resulted in a 30%–40% reduction in the risk of HHF and 40%–50% reduction in the risk of adverse renal outcomes among patients with diabetes with GFR ≧60 mL/min/1.73m^2^ (23). These findings support the benefits of SGLT2i for primary prevention in patients with type 2 diabetes but without overt ASCVD, HF, or CKD.

### Direct comparisons between dapagliflozin and empagliflozin

Given the concordant benefits of various SGLT2i, as well as their similar pharmacologic profiles, a class effect has been suggested for this drug entity. Although there have been some discrepancies in the reported clinical outcomes in clinical trials, this can be explained by differences in the inclusion criteria, baseline characteristics, and outcome definitions. However, several studies have suggested that SGLT2i effects may differ according to drug type. According to a multi-institutional cohort study by Shao et al., dapagliflozin may offer a more favorable benefit in terms of HF prevention, compared to that with empagliflozin (15). In another study by Shao et al., dapagliflozin had a more favorable HHF risk reduction than empagliflozin in patients with type 2 diabetes without ASCVD, while similar HHF risks were observed in patients with type 2 diabetes with ASCVD (16). The more potent effects of dapagliflozin over empagliflozin for HF outcomes can be explained by the SGLT2:SGLT1 receptor selectivity ratio, which is lower for dapagliflozin (1200:1) than for empagliflozin (2500:1) (24, 25). More specifically, myocardial ischemia and hypertrophy are associated with SGLT1 upregulation in the myocardium, where SGLT2 receptors are never expressed. This finding suggests that the SGLT2i with a lower specificity for SGLT2 receptors, and thus, a greater effect on SGLT1, has an even greater beneficial effect on HF prevention (26). In addition, compared to that with empagliflozin, dapagliflozin did not increase plasma aldosterone and noradrenaline levels, which could be advantageous for HF prevention (27).

In the present study, the benefits for HF and renal outcomes did not differ between dapagliflozin and empagliflozin groups. The changes in GFR also demonstrated similar patterns for these two SGLT2i, which showed significant benefits over DPP4i treatment (control group). Thus, in contrast to the studies by Shao et al.,(15, 16) we did not observe a more favorable benefit with dapagliflozin than with empagliflozin in terms of HF and renal outcomes. However, we acknowledge that the present study had a relatively small sample size and was performed in a retrospective manner. Although our findings support a SGLT2i class effect on HF and renal outcomes, future studies are warranted to compare their effects in a prospective trial or using nationwide real-world data.

Regarding the risk of ischemic events, a prospective observational study by Ku et al. found that empagliflozin resulted in better glycemic control and improvements in cardiometabolic components than that with dapagliflozin among patients with type 2 diabetes (17, 18). Similarly, in the present study, we found that HbA1c levels were significantly lower in the empagliflozin group than in the dapagliflozin and control groups. However, the LDLc levels at 24 and 36 months of treatment were higher in the empagliflozin group than in the other groups. It should be noted that better glycemic control, but higher LDLc level, with empagliflozin was not associated with clinical events; the risk of composite coronary and ischemic events was significantly lower in both SGLT2i groups than in the control group, without a significant difference between dapagliflozin and empagliflozin. Therefore, our study supports a SGLT2i class effect and, simultaneously, suggests that the benefits of SGLT2i use for ASCVD prevention are independent of the management of glucose and lipid profile. In particular, it has been suggested that SGLT2i can prevent endothelial dysfunction by reducing oxidative stress and sympathetic activation, reduce inflammatory cytokines, and reduce the plaque size while stabilizing their vulnerability (28).

### Study limitations

First, due to the relatively small sample size, the number of clinical events might not have been adequate to demonstrate a difference between dapagliflozin and empagliflozin. Second, laboratory tests were not performed with pre-specified schedules due to the retrospective nature of the study. However, more than 16,000 laboratory test results were included in the analysis, suggesting that our findings reflect real-world clinical practice. Third, although we used propensity-score matching to minimize differences in baseline characteristics between groups, some unmeasured confounders might remain unresolved.

## Conclusions

SGLT2i use was associated with a significantly reduced risk of ASCVD, HHF, and renal events among patients with diabetes and without prior ASCVD, HF, or CKD. There were no significant differences between dapagliflozin and empagliflozin, supporting a SGLT2i class effect.

## Data Availability

Data are available from the institutional review boards of Seoul National University Bundang Hospital (contact) for researchers who meet the criteria for access to confidential data. After ethical consideration in institutional review boards of Seoul National University Bundang Hospital, sharing a de-identified data set will be allowed.

## Acknowledgements

None.

